# BRAIN MR SPECTROSCOPIC FINDINGS IN THREE CONSECUTIVE COVID-19 PATIENTS: PRELIMINARY OBSERVATIONS

**DOI:** 10.1101/2020.06.10.20122465

**Authors:** Otto Rapalino, Akila Weerasekera, Sarah J. Moum, Katharina Eikermann-Haerter, Brian L. Edlow, David Fischer, Shibani Mukerji, Pamela W. Schaefer, R. Gilberto Gonzalez, Michael Lev, Eva-Maria Ratai

## Abstract

Brain magnetic resonance spectroscopic imaging (MRSI) was performed in three consecutive COVID-19 patients, as part of a pilot investigation of the pathophysiological processes underlying the brain involvement by the SARS-CoV-2 infection. These included one with necrotizing leukoencephalopathy, one after recent PEA cardiac arrest without leukoencephalopathy, and one without frank encephalopathy or recent severe hypoxic episode. The MRSI findings were compared to those of two patients with white matter pathology not SARS-CoV2 infection related, and a control patient without clinical encephalopathy. The N-acetylaspartate reduction, choline elevation, and glutamate/glutamine elevation found in the COVID necrotizing leukoencephalopathy patient and, to a lesser degree, the COVID post-cardiac arrest patient, follow a similar pattern as seen with the delayed post-hypoxic leukoencephalopathy patient. Lactate elevation was most pronounced in the patient with COVID necrotizing leukoencephalopathy.

## Introduction

Infection with severe Acute Respiratory Syndrome Coronavirus 2 (SARS-CoV-2) results in COVID-19 (Coronavirus Disease 2019), and declared a pandemic by the World Health Organization on March 11, 2020(1). As of early June 2020, millions of people worldwide have been sickened, and hundreds of thousands have died from this disease. COVID-19 primarily affects the lower respiratory tract, but many organs can be involved, including the central nervous system(2, 3). Neurological manifestations of SARS-CoV-2 infection are increasingly reported and include cerebrovascular complications, leukoencephalopathy, and other CNS disorders(4). Although COVID-19 associated white matter abnormalities resemble other forms of diffuse white matter injury (such as post-hypoxic leukoencephalopathy or sepsis-associated leukoencephalopathy), there are also notable differences, such as the neuroanatomical distribution of white matter lesions. The pathogenesis of COVID-19 white matter abnormalities remains unknown, although “silent hypoxia” has been hypothesized to have a role in its development(5).

While histopathology remains the reference standard for tissue characterization, MR spectroscopy represents a non-invasive *in-vivo* diagnostic tool for evaluating white matter injury and can provide valuable information regarding the underlying pathogenesis of white matter pathologies. In the setting of diffuse white matter injury, MRS offers a complementary evaluation to structural MR brain imaging by providing sensitive measurement of various *in-vivo* metabolites. Importantly, MRS can identify neurochemical abnormalities even in the absence of corresponding findings from MR brain imaging(6). Indeed, relatively specific patterns of metabolic derangements have already been established for several white matter pathologies(7).

The metabolic profile of COVID-19 associated leukoencephalopathy using MRSI has not been well established in the literature to date. We present examples of MR spectra in COVID-19 patients and compare them with both other leukoencephalopathy cases and a control case.

## Case series

Six MRSI datasets from three cases with COVID-19 **(Figure 1)**, two control leukoencephalopathy cases (post-hypoxic and sepsis-related leukoencephalopathy), and one control without encephalopathy were analyzed with LCModel, and their spectra were compared **(Table 1)**.

**Figure 1:**
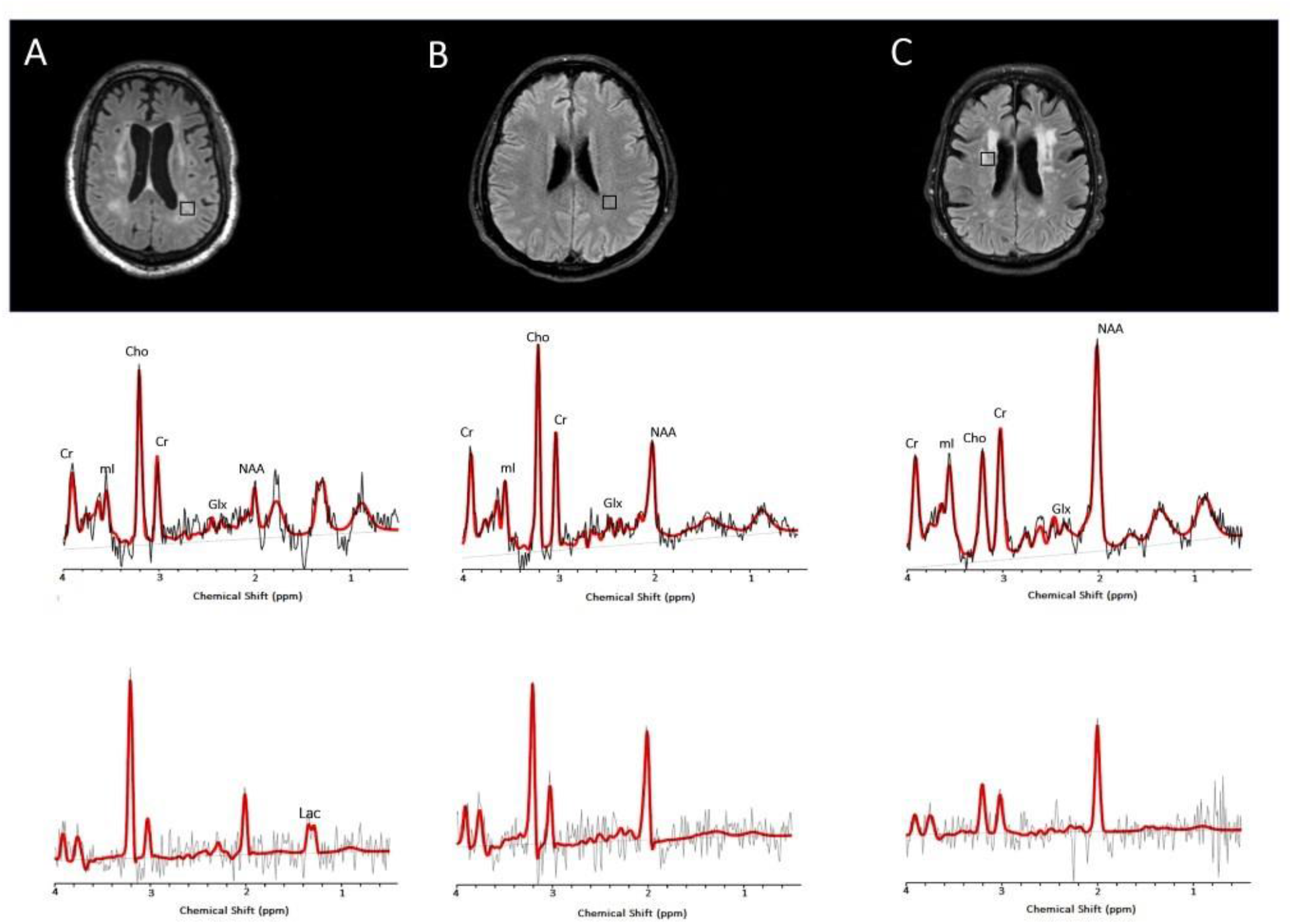
^1^H MR imaging spectra of three consecutive COVID-19 patients. **Upper panel** Axial T2/FLAIR images at the level of the corona radiata showing representative MRSI voxels (black squares), from hyperintense periventricular regions. **Lower panel:** Corresponding spectra (black) and LCModel fit (red) from each patient acquired at TE = 30ms (**top**) and TE = 288ms (**bottom**). (**A**) COVID-19 associated multifocal necrotizing leukoencephalopathy patient shows diffuse patchy white matter (WM) lesions with markedly increased choline (Cho) and decreased N-Acetyl-Aspartate (NAA), as well as elevated lactate (Lac). (**B**) COVID-19 patient after recent PEA cardiac arrest shows subtle FLAIR hyperintense lesions, also with markedly elevated Cho. Although the Cho/Cr ratio is also elevated, and NAA is decreased, these derangements are less severe than in patient A. There is no significant elevation of lactate. (**C**) COVID-19 patient without encephalopathy or recent severe hypoxia has normal Cho, with mildly decreased NAA and no elevation of lactate, from an anterior periventricular FLAIR hyperintense region.

**Table 1:** Demographics and clinical information.

| **Clinical**<br>**Diagnosis** | **Diagnosis** | **Status** | **Age** | **Gender** |
| --- | --- | --- | --- | --- |
| **COVID-A** | COVID-19 related<br>multifocal necrotizing<br>leukoencephalopathy | ICU and<br>intubated | 63 | M |
| **COVID-B** | COVID-19 related<br>recent PEA cardiac arrest,<br>without clear<br>leukoencephalopathy | ICU and<br>intubated | 53 | M |
| **COVID-C** | COVID-19 without clear<br>leukoencephalopathy or<br>recent severe hypoxia | Neurology ward<br>and non-<br>intubated | 72 | M |
| **DPL** | Non-COVID-19<br>Delayed post-hypoxic/toxic<br>leukoencephalopathy | Neurology ward<br>and non-<br>intubated | 44 | F |
| **SAE** | Non-COVID-19<br>Presumed sepsis-associated<br>encephalopathy | ICU and<br>mechanical<br>ventilation via<br>tracheostomy | 55 | F |
| **CONTROL** | Non-COVID-19 without<br>encephalopathy | Medicine ward<br>and non-<br>intubated | 55 | M |

Of the COVID-19 cases who had MRSI, one patient had MRI findings compatible with a necrotizing leukoencephalopathy with abnormal reduced diffusivity and cavitation within the white matter lesions (case COVID-A) **(Figure 1A**). The second patient had pulseless electrical activity (PEA) arrest while in the ICU and subsequently developed altered mental status. His MRI showed cerebellar cortical and hippocampal signal abnormalities suggestive of prior hypoxic-ischemic injury and subtle bilateral cerebellar and supratentorial white matter T2/FLAIR hyperintensities (COVID-B) (**Figure 1B**). The third patient without encephalopathic syndrome had a history of parkinsonism and developed catatonia of unclear etiology (COVID-C) (**Figure 1C**). This patient’s MRI showed mild nonspecific periventricular and deep white matter changes suggestive of small vessel disease (considering their neuroanatomical distribution).

These three cases were contrasted to MRSI data obtained prior to the COVID-19 pandemic from two patients with other leukoencephalopathies. One patient had delayed post-hypoxic/toxic leukoencephalopathy (DPL) and developed severe encephalopathy, rigidity, and mutism, likely related to a prior toxic exposure (opiates) or hypoxic episode (**Figure 2A**). The other non-COVID-19 patient had sepsis-associated encephalopathy (SAE) displaying mild diffuse white matter abnormalities on the brain MRI studies **(Figure 2B)**.

**Figure 2:**
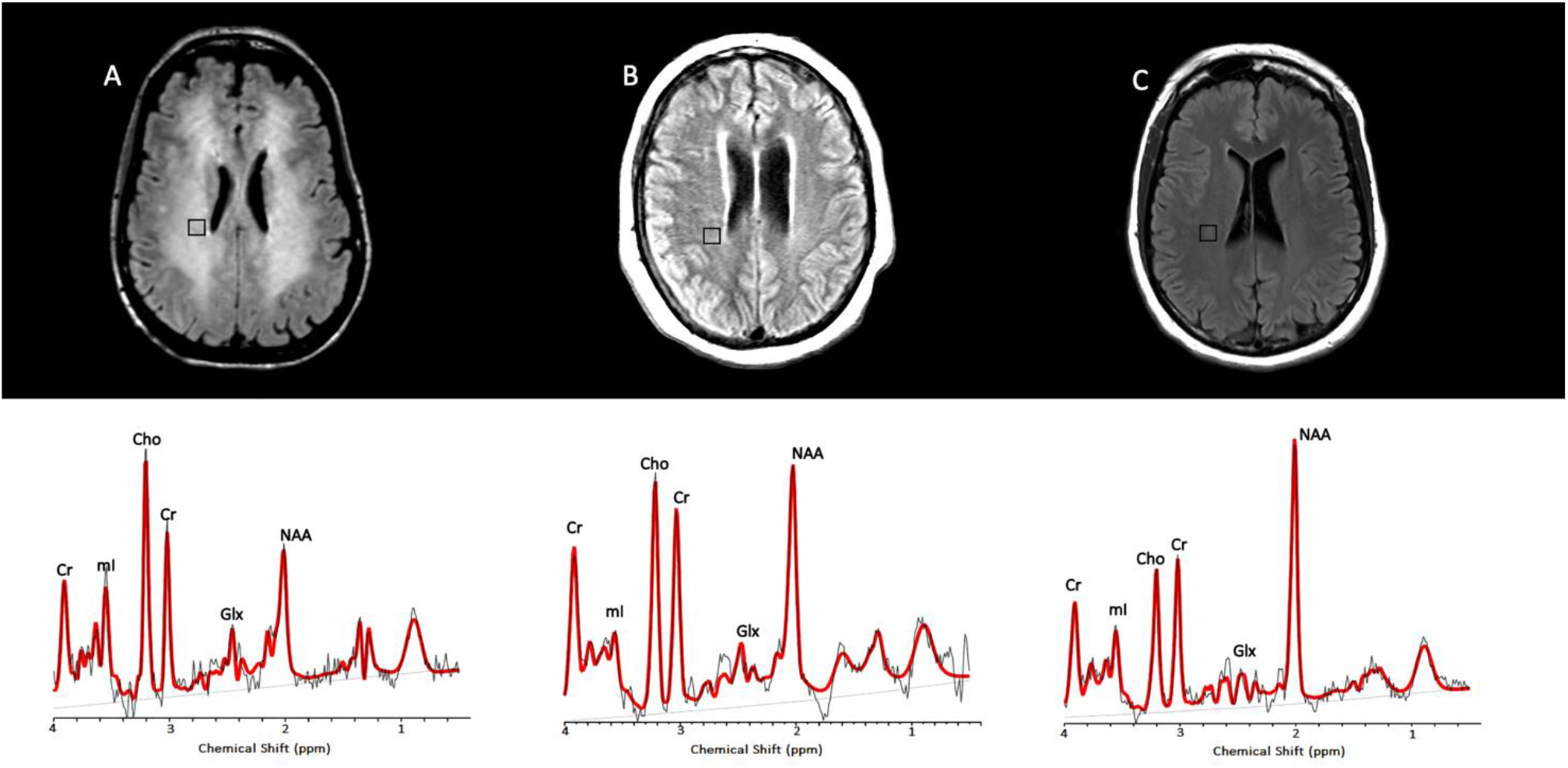
MRSI spectra from two non-COVID leukoencephalopathy patients, compared to a spectra from a control patient without leukoencephalopathy. Axial FLAIR/T2-weighted MR images and short TE MRSI spectra, obtained from areas of supratentorial signal abnormality with (**A**) delayed post-hypoxic/toxic leukoencephalopathy and (**B**) sepsis-associated encephalopathy, are compared to a corresponding region from a control patient (**C**) without encephalopathy.

Lastly, all five leukoencephalopathy patients were compared to the control case, a 55-year-old male presenting with paranoid delusions after acetaminophen overdose with a normal neurological exam and normal liver function tests at the time of the MRI. The control case had mild nonspecific white matter changes (**Figure 2C**).

MRSI datasets with short echo time (TE = 30ms) were analyzed to quantify ratios of NAA, Cho, mI, and Glx relative to creatine. Lac/Cr levels were quantified using MRSI datasets with long echo times(TE = 288ms). **Table 2** summarizes the means, standard deviations, and percent differences of metabolite levels to the control case.

**Table 2.** Mean, standard deviation, and percent difference relative to control, of brain MRSI metabolite ratios in three patients with and three patients without COVID-19.

| Diagnosis | No. Voxels | NAA/Cr* | Cho/Cr* | mI/Cr* | Glx/Cr* | Lac/Cr** |
| --- | --- | --- | --- | --- | --- | --- |
| **COVID-A** | 7 | 0.61±0.13<br>**(-96%)** | 0.65±0.05<br>**(+74%)** | 1.17±0.07<br>(+20%) | 1.46±0.20<br>**(+55%)** | 0.76±0.1 |
| **COVID-B** | 8 | 1.10±0.15<br>**(-44%)** | 0.54±0.04<br>**(+57%)** | 1.16±0.09<br>(+19%) | 1.40±0.13<br>**(+51%)** | 0 |
| **COVID-C** | 6 | 1.47±0.08<br>(-18%) | 0.27±0.03<br>(-10%) | 1.23±0.10<br>(+25%) | 1.04±0.22<br>(+24%) | 0 |
| **DPL** | 8 | 0.81±0.11<br>**(-72%)** | 0.48±0.02<br>**(+46%)** | 1.09±0.05<br>(+13%) | 1.82±0.14<br>**(+74%)** | - |
| **SAE** | 7 | 0.95±0.16<br>**(-58%)** | 0.32±0.04<br>(+6%) | 0.61±0.06<br>**(-45%)** | 1.10±0.12<br>(+28%) | 0 |
| **Control** | 7 | 1.73±0.08 | 0.30±0.03 | 0.96±0.09 | 0.83±0.14 | - |

(+/-%) indicate increase or decrease percent difference compared to the control patient; percent differences greater than 40% in magnitude are bolded. It is noteworthy that the distribution of bolded metabolite ratio derangements in COVID patients' **A** (leukoencephalopathy) and **B** (post cardiac arrest) are most like that of the delayed post-hypoxic/toxic leukoencephalopathy (**DPL**) patient.
\*: Data measured on short TE (TE=30 ms) spectra.
\*\*: Data measured on long TE (TE=288 ms) spectra. No long TE spectra were available for the DPL and control cases.

Comparative analysis showed decreased NAA/Cr levels within the white matter of all 3 COVID-19 positive patients compared to the control case. Of note, the patient with necrotizing leukoencephalopathy (COVID-A) displayed the most diminished NAA levels, which were completely absent in several MRS voxels; followed by the patient with recent cardiac arrest and mild diffuse white matter changes without necrosis (COVID-B); followed by the COVID-C case without cardiac arrest or overt leukoencephalopathy (**Figure 1**). The metabolic profiles of the 3 COVID-19 patients were contrasted to those of two non-COVID-19 patients with leukoencephalopathy (described above) (**Figure 2**). Their NAA/Cr levels were also markedly decreased but still higher than the COVID-A case (**Table 2)**.

Compared to the control case, two of the three COVID-19 patients had significantly elevated Cho/Cr levels (**Table 2**). The third COVID patient without leukoencephalopathy (COVID-C) showed Cho/Cr levels indistinguishable from those of the control case. The non-COVID-19 patient with DPL and significant white matter abnormalities showed increased Cho/Cr ratios, while the SAE patient displayed a relatively normal range of Cho/Cr.

All three COVID-19 patients and the non-COVID-19 patient with DPL showed slightly elevated myoinositol/Cr (mI/Cr) levels (**Table 2**). Interestingly, our patient with sepsis-associated encephalopathy had decreased levels of mI/Cr. Glutamate + glutamine/Cr was markedly increased in the two COVID-19 patients with leukoencephalopathy and after cardiac arrest as well as in the non-COVID-19 patient with DPL. Lac/Cr ratios were increased in the COVID-19 patient with necrotizing leukoencephalopathy on long TE spectra. No significant elevation of Lac/Cr ratios was seen in the SAE case. The DPL and control cases did not have long TE spectra, however, the short TE spectrum suggest the presence of lactate in the DPL case (**Figure 2A**).

## Discussion

Multiple recent reports in the medical literature have confirmed the development of leukoencephalopathy as a potential CNS complication in SARS-CoV-2 infection(8, 9), although no study to date has reported the MR spectroscopic findings in these patients. We evaluated the metabolic differences between patients with COVID-19 leukoencephalopathy, COVID-19 patients with relatively mild white matter involvement without a clear leukoencephalopathy, and other control groups to better understand the underlying pathophysiology of this disease. Markedly increased Cho/Cr, decreased NAA/Cr, and increased Lac/Cr ratios were observed in one COVID-19 patient with multifocal necrotizing leukoencephalopathy. Less pronounced changes in Cho/Cr and NAA/Cr ratios were noted in another COVID-19 case with prior PEA arrest and subtle nonspecific white matter signal abnormalities. Notably, the magnitude of the choline and NAA abnormalities in the patient with necrotizing leukoencephalopathy was more pronounced when compared to non-COVID-19 patients with delayed post-hypoxic leukoencephalopathy and sepsis-associated encephalopathy. In the setting of SARS-CoV-2 infection, the NAA/Cr ratios showed an inverse relationship, and the Cho/Cr ratios showed a positive relationship to the extent of observed white matter abnormalities.

In our series, similar patterns of metabolic changes were observed in the setting of COVID-19 associated leukoencephalopathy and delayed post-hypoxic leukoencephalopathy, namely elevated choline, elevated lactate, decreased N-acetyl-aspartate, increased myo-Inositol, and increased Glx ratios in relation to creatine. This spectral pattern has been previously reported in the setting of DPL following carbon monoxide poisoning or medication overdose(10–12). Prolonged impaired oxygenation to the subcortical white matter is thought to promote anaerobic metabolism and leads to elevated tissue lactate(11, 12).

Choline is a membrane marker in oligodendrocytes and astrocytes(13). The observed increase of Cho/Cr ratio in leukoencephalopathy cases, particularly in the case of COVID-19 associated necrotizing leukoencephalopathy, likely reflects demyelination within these white matter lesions as recently described in the first neuropathological study of a patient with COVID-19(14). Some choline elevation could be related to immune cellular infiltration within areas of demyelination(15). Axonal damage was also reported on pathology(14) and likely contributes to the decreased NAA/Cr ratios we observed, as NAA is considered a marker of neuronal and axonal integrity(16, 17).

Increased levels of myo-Inositol may reflect neuroinflammation, which when coupled with choline elevations in demyelinating pathologies may reflect glial proliferation(16). The presence of elevated Glx levels has been reported in cases of acute excitotoxic leukoencephalopathy(18) and viral-associated acute leukoencephalopathy with restricted diffusion (AESD)(19). Both conditions are thought to be mediated by excitotoxic injury to the cerebral white matter and exhibit prominent reduced diffusivity within white matter lesions.

Notably, the metabolic derangements seen in the setting of COVID-19 associated multifocal necrotizing leukoencephalopathy were more pronounced in our study than those observed with DPL. This finding raises the possibility of an additional or alternative underlying mechanism, other than the above-described hypoxia-driven process, contributing to the pathogenesis of COVID-19 associated leukoencephalopathy. Also, while based on a small sample size, our findings suggest that the metabolic changes in patients with SARS-CoV-2 infection increase with worsening white matter changes on conventional anatomic imaging. However, more evidence is required to validate this conclusion.

Our study’s small patient cohort limits our ability to generalize our observations. MRSI was only obtained in patients with SARS-CoV-2 infection when requested by referring providers for specific clinical indications.

In conclusion, the reported spectroscopic abnormalities within the white matter lesions of COVID-19 associated leukoencephalopathy may reflect several pathophysiological processes, including but not limited to: 1) an anaerobic metabolic environment produced by the well-described “silent hypoxia” seen in these patients resulting in elevation of lactate levels; 2) neuronal dysfunction and axonal injury with decreased NAA/Cr ratios; and, 3) increased membrane destruction or turnover with elevated Cho/Cr ratios. Other factors, in addition to hypoxia, might also be involved considering the magnitude of metabolite ratio changes we observed in these patients. Continued data collection in a larger cohort is required to validate these observations and better elucidate their significance in the pathophysiology of SARS-CoV-2 infection.

## Data Availability

Data generated or analyzed during the study are available from the corresponding author by request.

## ABBREVIATIONS

SARS-CoV-2: Severe Acute Respiratory Syndrome Coronavirus 2
COVID-19: Coronavirus Disease 2019
Cho/Cr: Choline/Creatine ratio
NAA: N-Acetyl-Aspartate
mI: Myo-Inositol
Lac: Lactate
Glx: Glutamate +
Glutamine: DPL = delayed post-hypoxic leukoencephalopathy
DPL: delayed post-hypoxic leukoencephalopathy
SAE: sepsis related encephalopathy
LCModel: Linear Combination of Model spectra
PEA: pulseless electrical activity

## Notes

Conflicts of Interest: There are no conflicts of interest relevant to this work.

### Competing Interest Statement

The authors have declared no competing interest.

### Funding Statement

None

### Author Declarations

This retrospective observational study was approved by our institutional review board (IRB no. 2015P001789) and conducted following Health Insurance Portability and Accountability Act (HIPAA) guidelines. The institutional review board waived written informed consent for this retrospective study.

## References

1. Cucinotta D, Vanelli M. WHO Declares COVID-19 a Pandemic. Acta Biomed 2020;91(1):157–160. doi: 10.23750/abm.v91i1.9397

2. Berlin DA, Gulick RM, Martinez FJ. Severe Covid-19. N Engl J Med 2020. doi: 10.1056/NEJMcp2009575

3. Romero-Sanchez CM, Diaz-Maroto I, Fernandez-Diaz E, Sanchez-Larsen A, Layos-Romero A, Garcia-Garcia J, Gonzalez E, Redondo-Penas I, Perona-Moratalla AB, Del Valle-Perez JA, Gracia-Gil J, Rojas-Bartolome L, Feria-Vilar I, Monteagudo M, Palao M, Palazon-Garcia E, Alcahut-Rodriguez C, Sopelana-Garay D, Moreno Y, Ahmad J, Segura T. Neurologic manifestations in hospitalized patients with COVID-19: The ALBACOVID registry. Neurology 2020. doi: 10.1212/WNL.0000000000009937

4. Helms J, Kremer S, Merdji H, Clere-Jehl R, Schenck M, Kummerlen C, Collange O, Boulay C, Fafi-Kremer S, Ohana M, Anheim M, Meziani F. Neurologic Features in Severe SARS-CoV-2 Infection. N Engl J Med 2020. doi: 10.1056/NEJMc2008597

5. Couzin-Frankel J. The mystery of the pandemic’s ‘happy hypoxia’. Science 2020;368(6490):455–456. doi: 10.1126/science.368.6490.455

6. Sun J, Song H, Yang Y, Zhang K, Gao X, Li X, Ni L, Lin P, Niu C. Metabolic changes in normal appearing white matter in multiple sclerosis patients using multivoxel magnetic resonance spectroscopy imaging. Medicine (Baltimore) 2017;96(14):e6534. doi: 10.1097/MD.0000000000006534

7. Bizzi A, Castelli G, Bugiani M, Barker PB, Herskovits EH, Danesi U, Erbetta A, Moroni I, Farina L, Uziel G. Classification of childhood white matter disorders using proton MR spectroscopic imaging. AJNR Am J Neuroradiol 2008;29(7):1270–1275. doi: 10.3174/ajnr.A1106

8. Radmanesh A, Derman A, Lui YW, Raz E, Loh JP, Hagiwara M, Borja MJ, Zan E, Fatterpekar GM. COVID-19 –associated Diffuse Leukoencephalopathy and Microhemorrhages. Radiology 2020:202040. doi: 10.1148/radiol.2020202040

9. Sachs JR, Gibbs KW, Swor DE, Sweeney AP, Williams DW, Burdette JH, West TG, Geer CP. COVID-19-Associated Leukoencephalopathy. Radiology 2020:201753. doi: 10.1148/radiol.2020201753

10. Beeskow AB, Oberstadt M, Saur D, Hoffmann KT, Lobsien D. Delayed Post-hypoxic Leukoencephalopathy (DPHL)-An Uncommon Variant of Hypoxic Brain Damage in Adults. Front Neurol 2018;9:708. doi: 10.3389/fneur.2018.00708

11. Shprecher DR, Flanigan KM, Smith AG, Smith SM, Schenkenberg T, Steffens J. Clinical and diagnostic features of delayed hypoxic leukoencephalopathy. J Neuropsychiatry Clin Neurosci 2008;20(4):473–477. doi: 10.1176/jnp.2008.20.4.473

12. Terajima K, Igarashi H, Hirose M, Matsuzawa H, Nishizawa M, Nakada T. Serial assessments of delayed encephalopathy after carbon monoxide poisoning using magnetic resonance spectroscopy and diffusion tensor imaging on 3.0T system. Eur Neurol 2008;59(1–2):55–61. doi: 10.1159/000109262

13. Urenjak J, Williams SR, Gadian DG, Noble M. Proton nuclear magnetic resonance spectroscopy unambiguously identifies different neural cell types. J Neurosci 1993;13(3):981–989.

14. Reichard RR, Kashani KB, Boire NA, Constantopoulos E, Guo Y, Lucchinetti CF. Neuropathology of COVID-19: a spectrum of vascular and acute disseminated encephalomyelitis (ADEM)-like pathology. Acta Neuropathol 2020. doi: 10.1007/s00401-020-02166-2

15. Brenner RE, Munro PM, Williams SC, Bell JD, Barker GJ, Hawkins CP, Landon DN, McDonald WI. The proton NMR spectrum in acute EAE: the significance of the change in the Cho:Cr ratio. Magn Reson Med 1993;29(6):737–745. doi: 10.1002/mrm.1910290605

16. Bitsch A, Bruhn H, Vougioukas V, Stringaris A, Lassmann H, Frahm J, Bruck W. Inflammatory CNS demyelination: histopathologic correlation with in vivo quantitative proton MR spectroscopy. AJNR Am J Neuroradiol 1999;20(9):1619–1627.

17. Soares DP, Law M. Magnetic resonance spectroscopy of the brain: review of metabolites and clinical applications. Clin Radiol 2009;64(1):12–21. doi: 10.1016/j.crad.2008.07.002

18. Takanashi JI, Murofushi Y, Hirai N, Sano K, Matsuo E, Saito K, Yasukawa K, Hamada H. Prognostic value of MR spectroscopy in patients with acute excitotoxic encephalopathy. J Neurol Sci 2020;408:116636. doi: 10.1016/j.jns.2019.116636

19. Kamate M. Acute Leukoencephalopathy with Restricted Diffusion. Indian J Crit Care Med 2018;22(7):519–523. doi: 10.4103/ijccm.IJCCM_139_18

